# A novel duplex qualitative real-time PCR assay for the detection and differentiation of Plasmodium ovale curtisi and Plasmodium ovale wallikeri malaria

**DOI:** 10.1101/2023.10.31.23297819

**Authors:** Wenqiao He, Rachel Sendor, Varun R. Potlapalli, Melchior M. Kashamuka, Antoinette K. Tshefu, Fernandine Phanzu, Albert Kalonji, Billy Ngasala, Kyaw Lay Thwai, Jonathan J. Juliano, Jessica T. Lin, Jonathan B. Parr

## Abstract

**Background:** *P. ovale* spp. infections are endemic across multiple African countries and are caused by two distinct non-recombining species, *P. ovale curtisi* (*Poc*) and *P. ovale wallikeri* (*Pow*). These species are thought to differ in clinical symptomatology and latency, but existing diagnostic assays have limited ability to detect and distinguish them. In this study, we developed a new duplex assay for the detection and differentiation of *Poc* and *Pow* that can be used to improve our understanding of these parasites.

**Methods:** Repetitive sequence motifs were identified in available *Poc* and *Pow* genomes and used for assay development and validation. We evaluated the analytical sensitivity and specificity of the best-performing assay using a panel of samples from Tanzania and the Democratic Republic of the Congo (DRC), then validated its performance using 55 *P. ovale* spp. samples and 40 non-ovale *Plasmodium* samples from the DRC. *Poc* and *Pow* prevalence among symptomatic individuals sampled across three provinces of the DRC were estimated.

**Results:** The best-performing *Poc* and *Pow* targets had 9 and 8 copies within the reference genomes, respectively. Our duplex assay had 100% specificity and 95% confidence lower limits of detection of 4.2 and 41.2 parasite genome equivalents/µl for *Poc* and *Pow*, respectively. Species was determined in 80% of all *P. ovale* spp.-positive field samples and 100% of those with >10 parasites/µl. Most *P. ovale* spp. field samples from the DRC were found to be *Poc* infections.

**Conclusions:** We identified promising multi-copy targets for molecular detection and differentiation of *Poc* and *Pow* and used them to develop a new duplex real-time PCR assay that performed well when applied to diverse field samples. Though low-density *Pow* infections are not reliably detected, the assay is highly specific and can be used for high-throughput studies of *P. ovale* spp. epidemiology among symptomatic cases in malaria-endemic countries like the DRC.

**Author Summary:** Non-falciparum malaria is gaining attention, especially in settings where *P. falciparum* transmission is declining. *Plasmodium ovale curtisi* (*Poc*) and *wallikeri* (*Pow*) are neglected parasites that can cause relapsing malaria and are thought to differ in clinical symptomatology and latency. However, existing diagnostic assays have limited ability to detect and distinguish *Poc* and *Pow* and are not well-suited for high-throughput use, hindering our understanding of *P. ovale* spp. epidemiology. Mining recently available *Poc* and *Pow* reference genomes, we identify new multi-copy targets for molecular detection and develop a novel duplex qualitative real-time PCR assay capable of species differentiation. The assay is highly specific and requires short turn-around time. While sensitivity can be improved for low-density *Pow* infections, this new assay can be used for high-throughput studies of symptomatic *P. ovale* spp. infections in malaria-endemic countries. We apply this tool to samples collected during a large study conducted in the DRC and investigate *P. ovale* spp. epidemiology across health centers in three provinces.

## Introduction

Malaria remains a major global health concern despite decades of sustained investment in elimination efforts. Though most malaria programs prioritize *Plasmodium falciparum*, the parasite species responsible for most deaths, increasing evidence confirms co-circulation of other neglected *Plasmodium* species that cause human malaria [1]. Recent surveys reveal a previously unappreciated burden of *Plasmodium ovale* spp. in multiple African countries [2, 3], where relapsing malaria caused by *P. ovale* spp. may prove to be an obstacle to malaria elimination efforts [4–6]. *P. ovale* comprises two distinct non-recombining species, *P. ovale curtisi* (*Poc*) and *P. ovale wallikeri* (*Pow*) [7]. *Poc* and *Pow* have potential differences in clinical symptomatology and latency [8], but existing diagnostic assays have limited ability to detect and distinguish them, and require multiple steps or prolonged cycling time that increases risk of false-positive results [9].

Detection and differentiation of *Poc* and *Pow* is not currently possible using conventional malaria diagnostic assays relying on microscopy. Microscopic examination of blood smears remains the gold standard for malaria diagnosis, but differentiation of parasite species and examination of low parasite density or mixed-species infections is challenging and requires significant expertise [10]. *Poc* and *Pow* infections often occur as mixed infections at low density, and are morphologically indistinguishable on blood slides [11, 12]. Furthermore, widely used malaria rapid diagnostic tests (RDTs) fail to detect samples with low parasite densities and cannot distinguish parasite species other than *P. falciparum* and *Plasmodium vivax* [13, 14]. Thus, alternative methods are required to identify these neglected species.

Molecular methods are more sensitive and specific for *Plasmodium* detection than microscopic examination or RDTs, but most existing assays target the 18S rRNA gene of both *P. ovale* spp. leading to potential cross-reactivity and a lack of complete species specificity for *Poc* and *Pow*. A duplex real-time PCR assay for *Poc* and *Pow* detection was published in 2011, however, results of the melt-curve analysis can be hard to interpret [15]. A nested PCR assay was developed in 2013 [16], and while it can detect samples with 2-10 parasites/µl, this assay requires multiple steps and long turnaround time. Available single-target quantitative real-time PCR assays require separate runs to distinguish *Poc* and *Pow* [9, 17, 18]. Because of the limitations of the existing assays, most studies of *P. ovale* spp. have not distinguished *Poc* and *Pow.* However, recently released *Poc* and *Pow* genomes (PocGH01 and PowCR01) provide opportunities for improved molecular assay development [19]. To improve our understanding of the epidemiology of *P. ovale* spp. malaria, we mined publicly available *Poc* and *Pow* genomes to identify novel multi-copy targets and developed a new duplex qualitative real-time PCR assay for detection and differentiation of *Poc* and *Pow* infection.

## Materials and Methods

### Mining and selection of multi-copy targets in *P. ovale curtisi* and *P. ovale wallikeri* genomes

Using the publicly available *Poc* (PocGH01) and *Pow* (PowCR01) reference genomes obtained from the NIH National Center for Biotechnology Information (NCBI) database, we identified sequence motifs of 100 base-pairs (bp) in length with ≥6 copies using *Jellyfish* (version 2.2.10) [20] (Fig 1). Sequences with low GC content (< 25%) and highly repetitive short sequences were excluded. The remaining multi-copy targets were aligned to NCBI nt database using *blastn* to investigate their specificity. Sequences aligned to other *Plasmodium* parasites were excluded. We then re-aligned the remaining targets to the *Poc* and *Pow* genomes separately using *blastn* to investigate their copy numbers in each genome. Candidate diagnostic assay targets for *Poc* and *Pow* were selected based on species-specificity and copy numbers. Primer and probe sets were designed manually using Oligo Calc [21] and DNAMAN (version 9, Lynnon BioSoft, Quebec City, Canada) to estimate primer and probe melting temperatures and to avoid self-complementarity and primer dimers (S1 Table).

**Fig 1.**
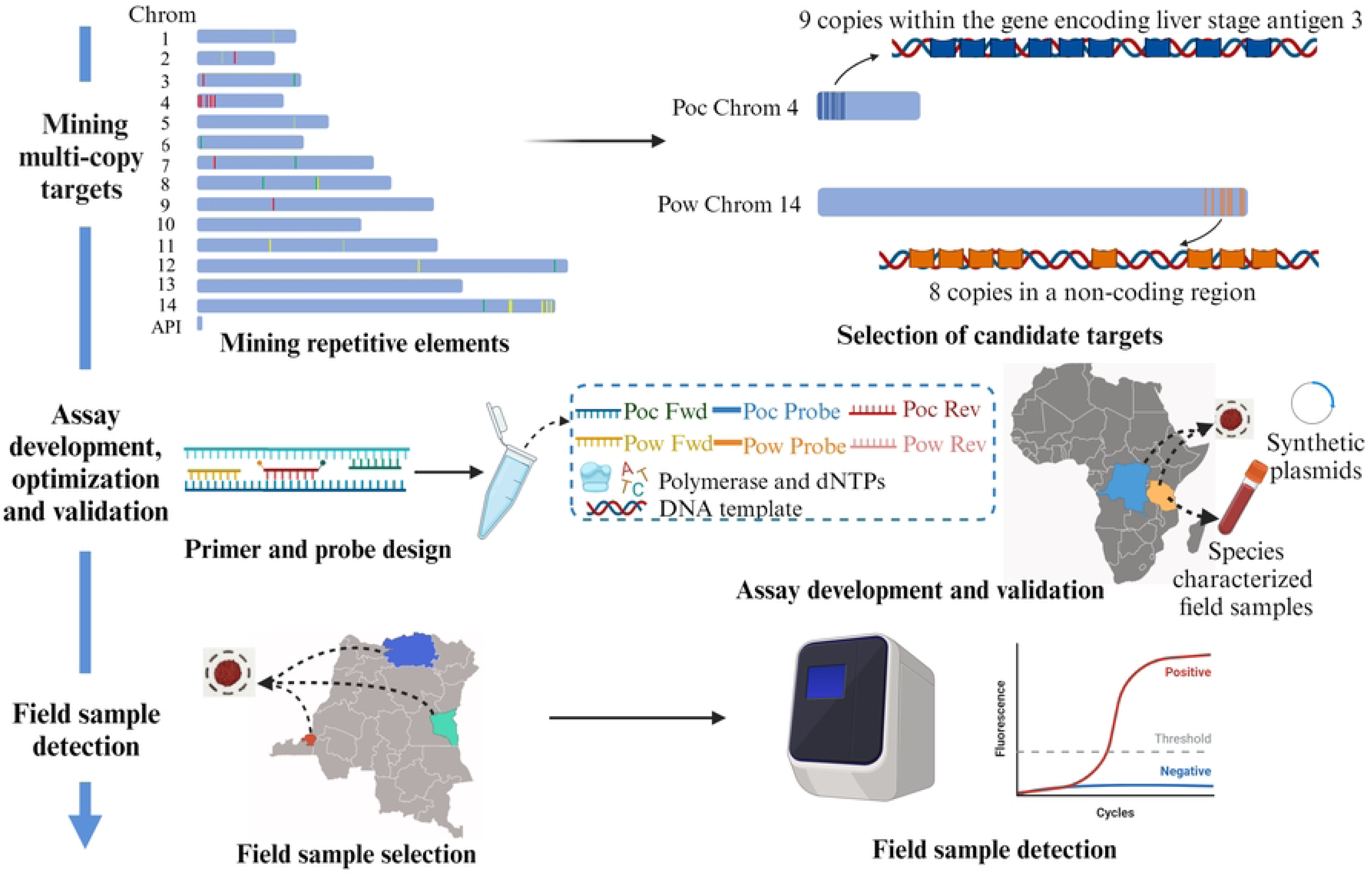
Approach to developing a real-time PCR assay for detection and differentiation of *P. ovale curtisi* and *P. ovale wallikeri*.

### Assay development and optimization

A panel of 15 well-characterized *Poc* and *Pow* field samples and six non-ovale *Plasmodium* laboratory controls were selected for assay development and analytical specificity analysis. Field samples included 11 *Poc* and four *Pow* leukodepleted blood samples and dried blood spot (DBS) samples from Tanzania and the Democratic Republic of the Congo (DRC). Laboratory controls included two *P. falciparum*, one *P. malariae*, two *P. vivax*, and one *P. knowlesi* samples from an external quality assurance program [22]. DNA was extracted from dried blood spot (DBS) and leukodepleted blood samples using Chelex 100 (Bio-Rad, Fishers, Indiana, USA) [23] and the QIAamp DNA Mini Kit (Qiagen, Mettmann, North Rhine-Westphalia, Germany), respectively. *P. ovale* parasite densities were estimated using a semi-quantitative real-time PCR assay targeting the 18S rRNA gene of both *P. ovale* spp. as previously described [2]. *Poc* versus *Pow* species was determined using two singleplex real-time PCR assays as previously described [9].

Primer sets with the best specificity for *Poc* and *Pow* versus this panel of samples were selected. A duplex qualitative real-time PCR assay was developed based on the selected primer sets and the corresponding probes. Assay optimization was then performed using synthetic plasmids containing targets for *Poc* and *Pow* detection (Azenta Life Sciences, Indianapolis, Indiana, USA). We tested a range of annealing temperatures and of primer and probe concentrations to identify optimal reaction conditions. All optimization analyses were performed in duplicate.

### Analytical sensitivity and specificity

We further characterized the best performing assay to determine its analytical sensitivity and specificity. Analytical sensitivity estimates were determined using probit analysis [24] with serially diluted *Poc* and *Pow* plasmid DNA comprising 104 and 161 total replicates, respectively (S2 Table). Analytical specificity of the present assay was assessed using the same panel of 15 well-characterized *Poc* and *Pow* field samples and six non-ovale *Plasmodium* laboratory controls described above in duplicate. Samples with Ct values less than 45 in both reactions were considered positive.

### Validation using field samples

The assay’s clinical sensitivity and specificity were assessed using 95 dried blood spot samples selected from a large sample set from a previous study conducted in the DRC [25], including 55 *P. ovale* spp. samples and 40 non-ovale *Plasmodium* samples (20 *P. falciparum* infections, 10 *P. malariae* infections, and 10 *P. falciparum* and *P. malariae* mixed infections) [25]. DNA was extracted using Chelex 100 as described above. *Plasmodium species* and parasite densities were identified using real-time PCR assays for *P. ovale*, *P. falciparum* and *P. malariae* as previously described, with samples positive in duplicate selected for use during validation of the present assay, and the previously published singleplex real-time PCR assay for both *P. ovale* spp. used as the gold standard for clinical sensitivity and specificity calculations [2, 26, 27]. Samples with Ct values lower than 45 in duplicate by our new assay were considered to be positive.

### Epidemiology of *Poc* and *Pow* among symptomatic patients in the DRC

We investigated the epidemiology of *P. ovale* spp. infection and the distribution of *Poc* vs *Pow* infections using samples from a study of symptomatic malaria across three provinces in the DRC. Among a randomly selected group of 1,000 symptomatic individuals, 64 samples were previously found be positive for *P. ovale* spp. by real-time PCR testing [2, 25]. The χ^2^ test or Fisher’s exact test was used for comparison with categorical variables that might be associated with *P. ovale* spp.-infections based on results of previously published studies [28–33], including age, sex, *P. falciparum* co-infection, bed net use, education level, recurrent malaria infection (any prior *Plasmodium* species infections within 6 months), and rural residence. Though no significant association between pregnancy and malaria was reported in previous studies, pregnancy was included in the present analysis.

We calculated crude odds ratios (cORs) and their 95% confidence intervals (CI) to evaluate associations between each of these eight factors and *P. ovale* infections. Because DNA degradation is possible during long-term storage, we repeated the previously published singleplex *P. ovale* spp. real-time PCR assay on the 64 samples previously identified as *P. ovale* spp.-positive. Among these, 44 tested positive in duplicate and were selected for species determination using the present duplex assay. To determine the proportions of *Poc* and *Pow* within these *P. ovale* spp.-positive samples, inverse probability weighting (IPW) was used to account for differences between selected samples and the original 64 *P. ovale* spp.-positive samples. Selection weights were calculated by multiple logistic regression analysis with the following covariates: gender, age, area, and *P. ovale* spp. parasitemia.

### Statistical analysis

Statistical analysis was performed using R software (version 4.2.0; R Core Team, Vienna, Austria) in RStudio (version 2022.02.2). Figures were generated using the *ggplot2* (version 3.4.1) and *forestplot* (version 3.1.1) packages. Mapping was done via pixelmap [34].

### Ethical approvals

Existing samples from previous studies were chosen based on convenience. DRC samples were collected as part of a 2017 study investigating malaria diagnostic test performance in three provinces, Kinshasa, Bas-Uele, and Sud-Kivu[25]. Tanzania samples were collected from participants enrolled in a malaria transmission study in rural Bagamoyo district from 2018-2019 [9, 35, 36]. Enrolled subjects provided informed consent or assent. Ethical approvals for these studies were obtained from the Kinshasa School of Public Health, Muhimbili University of Health and Allied Sciences, and the University of North Carolina at Chapel Hill.

## Results

### *P. ovale curtisi* and *P. ovale wallikeri* target selection and assay development

A total of 2,585 and 3,978 sequences of 100 bp in length with ≥ 6 repeats were found in the *Poc* and *Pow* reference genomes, respectively. Targets with low GC content, highly repetitive short sequences, or aligned to other *Plasmodium* parasite genomes were excluded. A total of three potential assay targets with ≥8 copies in each of the *Poc* and *Pow* genomes were selected. Focusing on these potential targets, we designed five and three primer and probe sets for *Poc* and *Pow*, respectively (S1 Table). After testing all primer and probe sets using a panel of 15 well-characterized *Poc* and *Pow* field samples and six laboratory non-*ovale Plasmodium* controls, we selected two primer and probe sets with the best specificity for *Poc* and *Pow*, respectively, for additional laboratory testing (Table 1). The selected *Poc* target had nine copies within putative liver stage antigen 3 (*lsa3*) gene on chromosome 4 (LT594585.1: 9,968-11,125), while the *Pow* target had eight copies in a non-coding region on chromosome 14 (LT594518.1: 1,842,975-1,844,586). Short distances (< 50 bp) were noted between the repetitive *Poc* target motifs and between *Pow* target motifs.

**Table 1.**
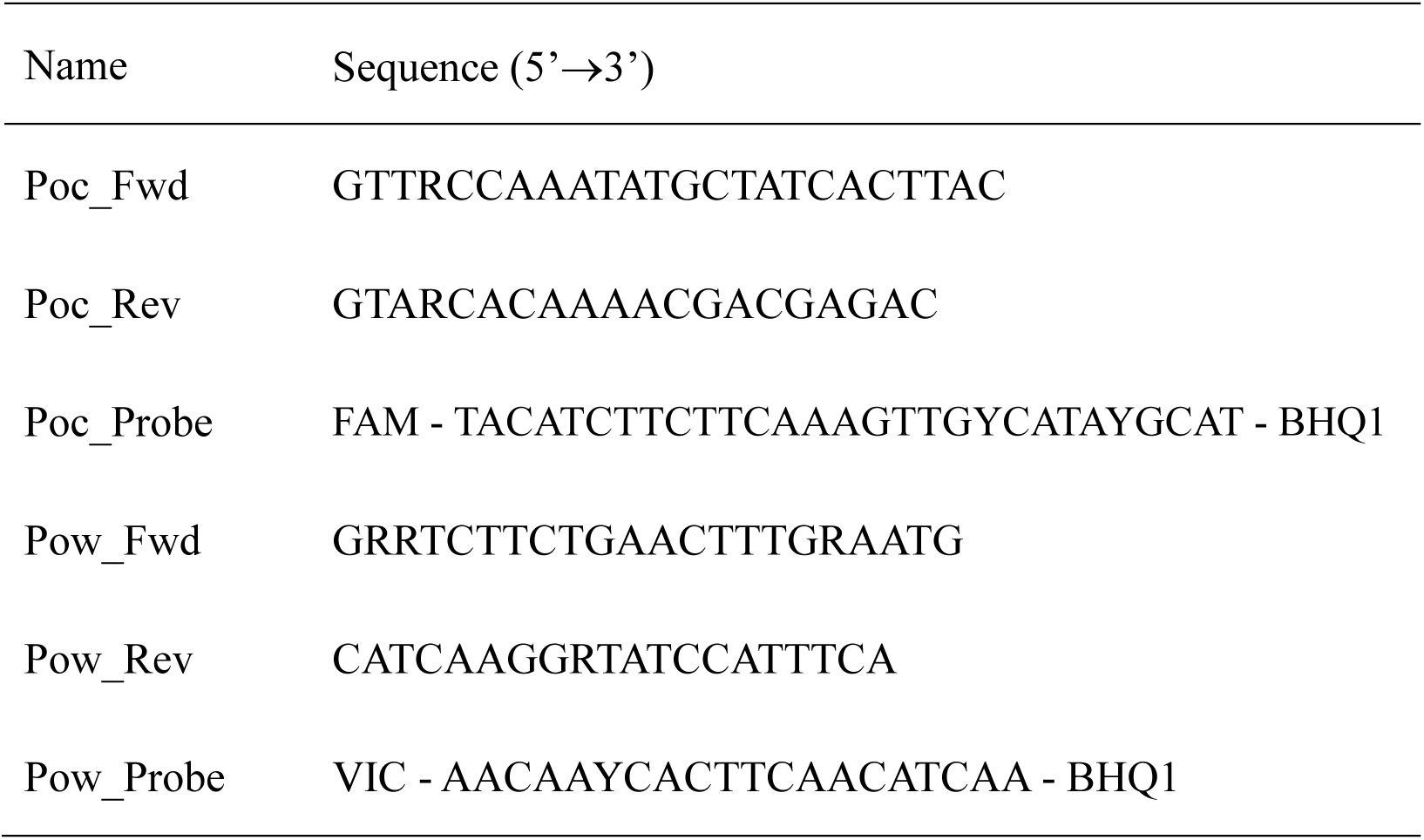
Best performing primers and probes for *P. ovale curtisi* and *P. ovale wallikeri* detection.

Combining these *Poc* and *Pow* primers and probes, we optimized a duplex, qualitative real-time PCR assay for the detection and differentiation for *Poc* and *Pow*. The final, optimized duplex assay was performed in a small final volume of 10µl, including 7µl of reaction master-mix containing 2x FastStart Universal Probe Master (Rox) (Roche, Basel, Switzerland), primers and probes (240 nM of Poc_Fwd, 240 nM of Poc_Rev, 60 nM of Poc_Probe, 800 nM of Pow_Fwd, 800 nM of Pow_Rev, 320nM of Pow_Probe); and 3µl of DNA template. Optimal thermocycling conditions were two min at 50°C, 10 min at 95°C, followed by 45 cycles of 15 s at 95°C and 60 s at 58°C, allowing for detection of parasite DNA in less than two hours.

### Analytical sensitivity and specificity

The 95% confidence lower limits of detection for *Poc* and *Pow* were 4.2 and 41.2 parasite genome equivalents/µl, respectively (Fig 2A, S1 Fig and S2 Table). All well-characterized *Poc* and *Pow* field samples were successfully detected and differentiated with no cross-reactivity between species, and no cross reactivity was found in six non-ovale *Plasmodium* controls (Fig 2B), consistent with 100% analytical specificity.

**Fig 2.**
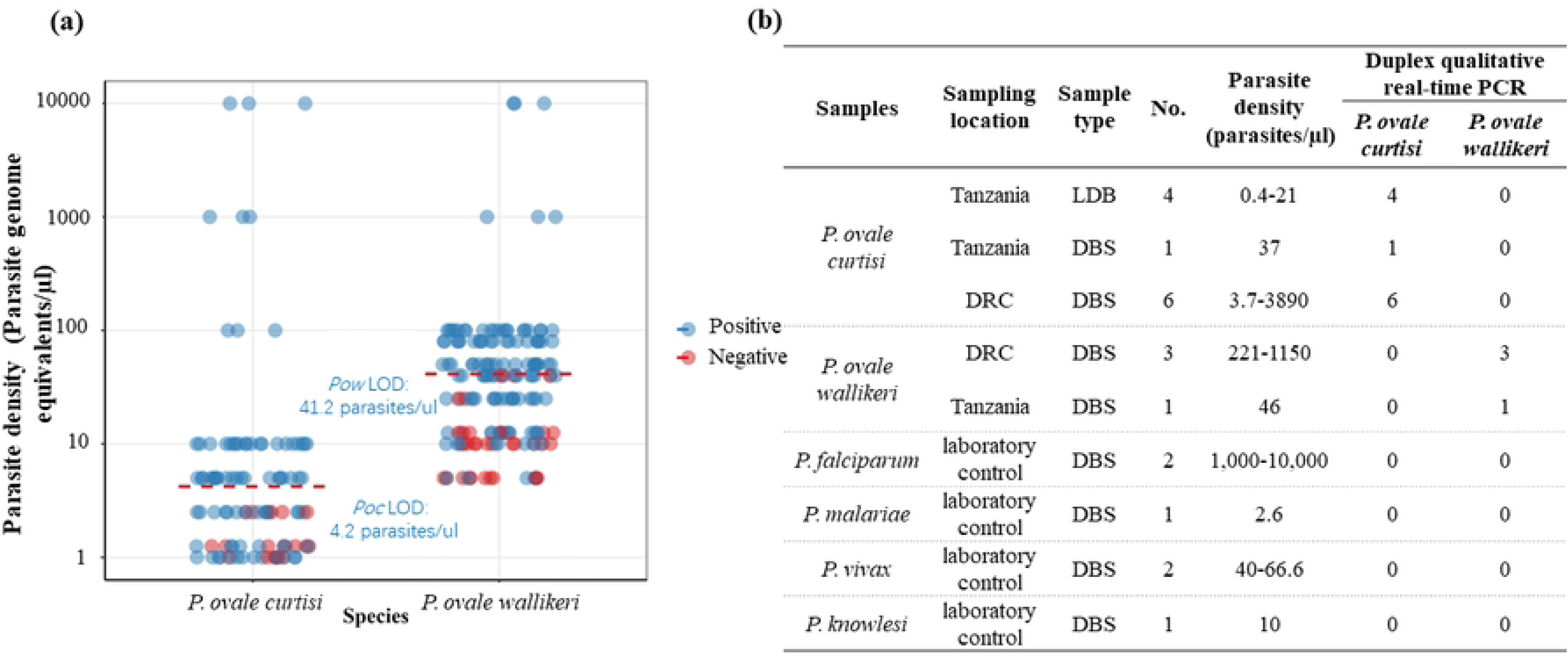
Duplex *Poc* and *Pow* assay performance. **A)** Analytical sensitivity when applied to multiple replicates of serially diluted plasmid DNA (n=104 and 161 total replicates for *Poc* and *Pow*, respectively). Points are colored to display target detection (blue) versus no detection (red). The 95% lower limit of detection (LOD) determined using probit analysis is shown for each species. **B)** Analytical specificity versus genomic DNA extracted from a panel of well-characterized leukodepleted blood (LDB) and dried blood spot (DBS) samples from Tanzania and the DRC with *Poc* and *Pow* confirmed by nested PCR, and non-ovale *Plasmodium* samples from an external quality assurance program. All *Poc* and *Pow* samples were correctly identified, and no false-positives were observed among other *Plasmodium* species.

### Validation using field samples

The assay demonstrated excellent clinical sensitivity at higher parasite densities and perfect specificity when applied to 95 field samples collected in the DRC. Parasite densities of 55 *P. ovale* spp.-positive field samples included in this study ranged from 0.9 parasites/µl to 2,468 parasites/µl; 29 (52.7%) had parasite densities <10 parasites/µl. The assay’s overall sensitivity was 80%, successfully determining *P. ovale* species in 44 of the *P. ovale* spp.-positive field samples (Fig 3A); however, assay sensitivity was 100% for infections with >10 parasites/µl. The lowest parasite densities in which species could be determined were 2.0 and 20.9 parasites/µl for *Poc* and *Pow*, respectively. None of the 40 non-ovale *Plasmodium* field samples were detected by the duplex assay, consistent with 100% specificity (Fig 3B).

**Fig 3.**
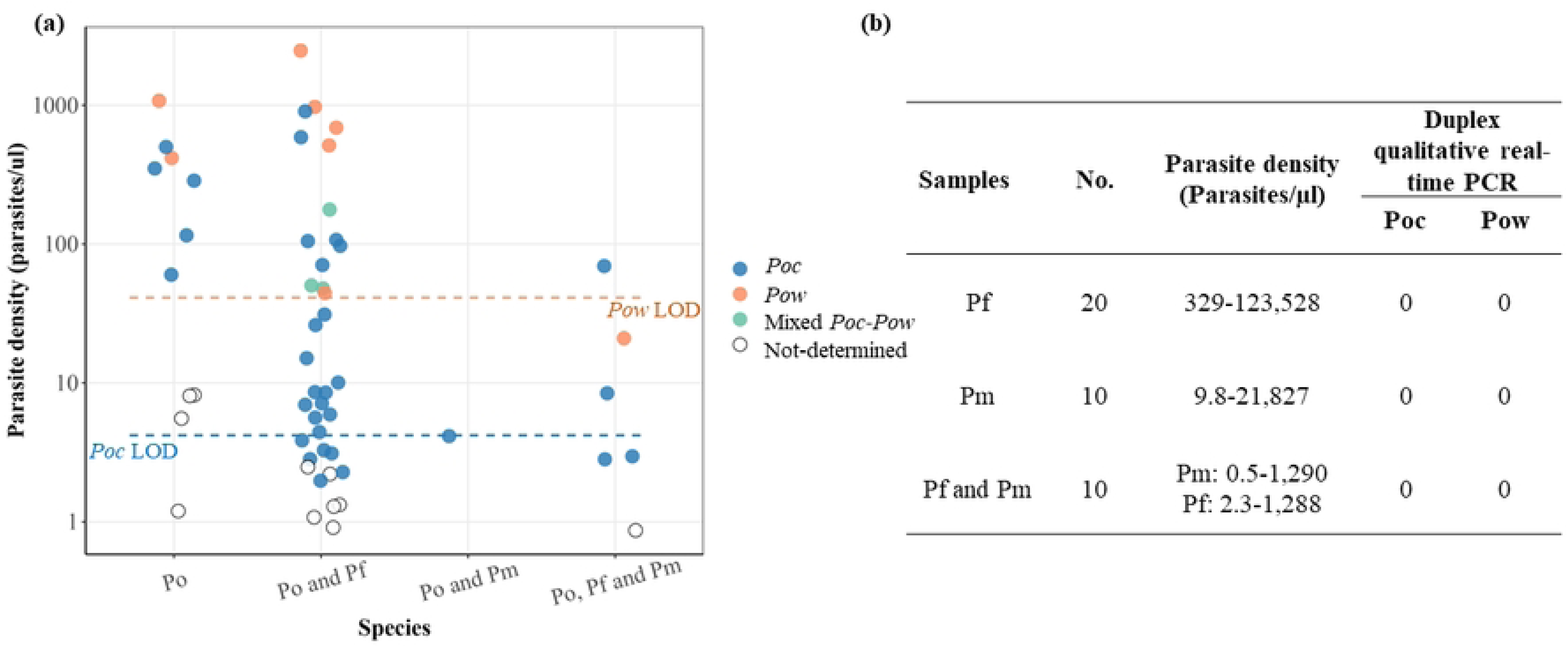
Assay validation using field samples collected in the DRC. Gold standard species identification was performed previously using a series of semi-quantitative qPCR assays targeting pan-*Plasmodium* 18S rRNA, followed by singleplex species-specific assays including *P. ovale* spp. [2] **A)** Detection of known *P. ovale* spp. PCR-positive samples with varying parasite densities and co-infection status. Analytical 95% lower limits of detection (LOD) are represented by dashed lines. **B)** No detection of other *Plasmodium* species across a range of parasite densities. Abbreviations: Po = *P. ovale* spp. (comprising both *Poc* and *Pow*); *Poc* = *P. ovale curtisi*; *Pow* = *P. ovale wallikeri;* Pf = *P. falciparum*; *Pm = P. malariae*.

### Epidemiology of symptomatic malaria due to *P. ovale* in the DRC

A total of 64 *P. ovale* spp.-positive samples were previously identified among randomly selected field samples from 1,000 participants in three provinces in the DRC [25]. The prevalences of *P. ovale* spp.-infection in Bas-Uele, Kinshasa, and Sud-Kivu were 14.3% (47/328), 2.8% (10/353), and 2.2% (7/319), respectively. Among these 1,000 participants, urban residence (cOR: 0.31, 0.17-0.55) and bed net use (cOR: 0.50, 0.24-0.85) had significant protective associations with *P. ovale* spp. infection; while recurrent malaria infection (within 6 months) (cOR: 1.75, 1.02-3.03) and coinfection with *P. falciparum* (cOR: 7.70, 3.48-17.06) was associated with *P. ovale* spp. infection (Fig 4).

**Fig 4.**
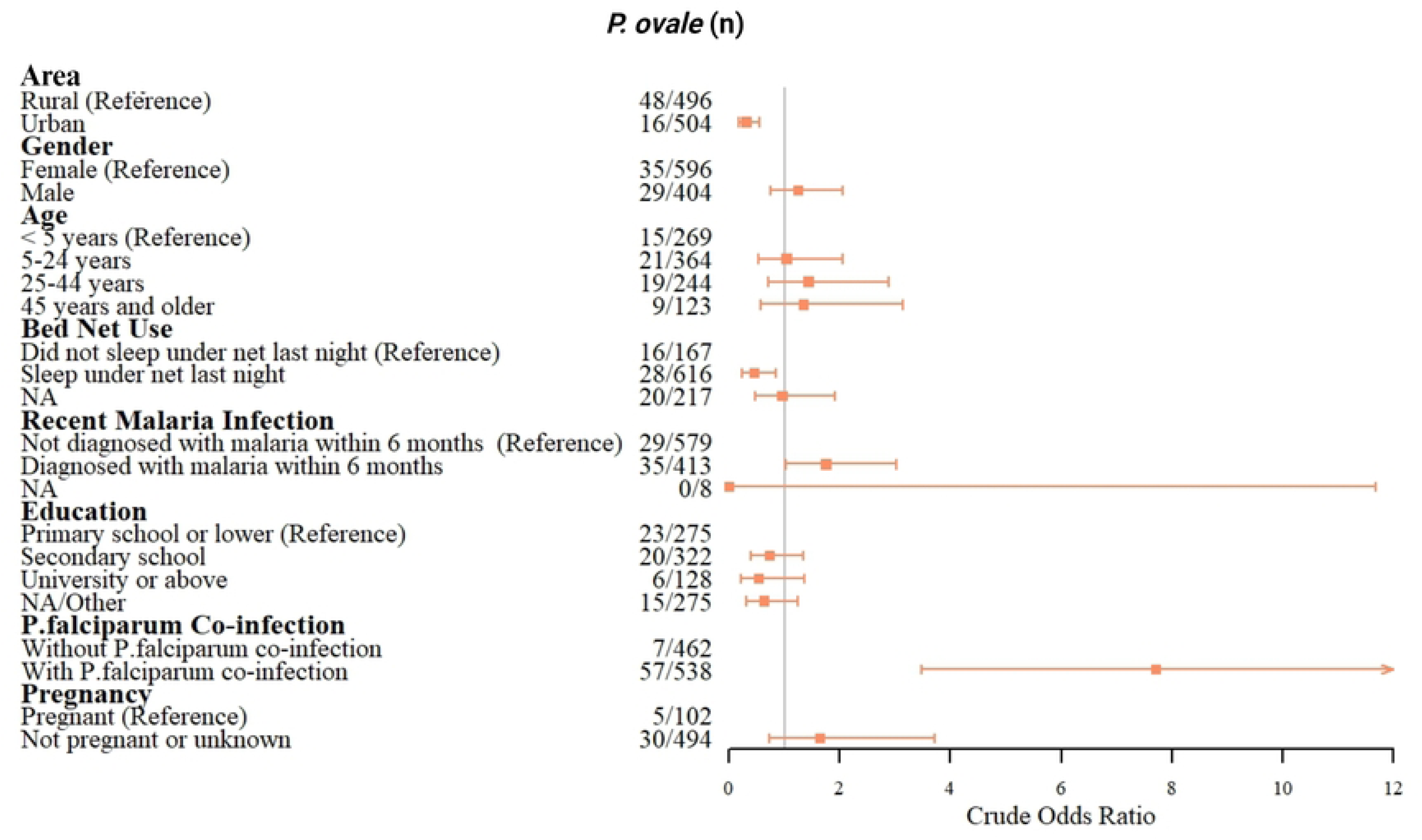
Factors associated with *P. ovale* spp. (comprising both *Poc* and *Pow*) infections. Crude odds ratio estimates and associated 95% confidence intervals are displayed.

Among the subsample of 44 *P. ovale* spp.-positive samples selected for species determination, *P. ovale* species was determined in 37 (84%) samples using the present assay: 28 (75.7%), 6 (16.2%), and 3 (8.1%) were *Poc* infections, *Pow* infections, and *Poc*-*Pow* mixed infections, respectively. Inverse probability weighting analysis indicated that 89.8% of all *P. ovale* spp. infections in our cohort included *Poc* species, alone or as a *Poc*-*Pow* mixed infection, and 15.2% included *Pow* species, alone or as a *Poc*-*Pow* mixed infection.

## Discussion

We mined recently published genomes of *Poc* and *Pow* to develop a highly specific duplex real-time PCR assay that can be used to improve our understanding of their epidemiology in malaria-endemic countries. Recent studies have revealed a previously unappreciated burden of *P. ovale* spp. in Africa [2, 15, 36, 37]. Though *Poc* and *Pow* are distinct non-recombining species, the few existing assays capable of distinguishing them are not well-suited to large studies, requiring separate assays for each species and higher volumes of DNA, with potential for cross-reactivity at higher parasite densities. Because current assays require multiple steps, long turnaround time, or lack complete species specificity [15–18], most field studies do not distinguish *Poc* and *Pow*, and their prevalence and clinical features remain understudied [38–40]. Our new assay can be used to help bridge this knowledge gap, especially among symptomatic cohorts where low parasite density infections are not predominant.

Our assay targets are distinct from those used in prior assays and take advantage of 100 bp repetitive motifs in the putative *lsa3* gene on *Poc* chromosome 4 and a non-coding region on *Pow* chromosome 14, respectively. Studies of *P. falciparum lsa3* indicate that it is a mutable non-essential gene expressed during the pre-erythrocytic stages of infection that encodes an antigen with tetrapeptide repeats of unclear function [41, 42]. *P. falciparum lsa3* was shown to be largely conserved across isolates collected from geographically diverse sites [42]. The non-coding *Pow* repetitive motif we targeted has unclear function, with no obvious orthologues identified in publicly available databases. Though non-essential genes are more likely to be lost over time based on *in vitro P. falciparum* studies, genes with putative roles in antigenic variation such as *lsa3* might be important in human infection [41–44]. We leveraged the repetitive nature of these poorly understood *Poc* and *Pow* targets to develop an assay with several advantageous performance characteristics.

Compared to published real-time PCR assays that usually target *Poc* and *Pow* 18S rRNA genes [2, 17, 18], inclusion of distinct *Poc* and *Pow* targets enabled development of a highly specific assay. These targets’ copy numbers are higher than those reported for 18S rRNA genes in *Plasmodium* genomes [45, 46]. However, published 18S rRNA PCR assays achieve similar limits of detection compared to our assay, with 1.5 parasites/µl and 50 plasmid copies/µl for *Poc* and *Pow* detection, respectively [17, 18]. It is possible that the short distances between our *Poc* targets and between *Pow* targets decrease the PCR efficiency, offsetting improved sensitivity that might otherwise be achieved from their increased copy number. Validation using field samples from the DRC confirmed robust species differentiation when the assay was applied to *P. ovale* spp. samples with >10 parasites/µL and 100% specificity across all parasite densities. Though its ability to identify *Pow* in particular was limited at lower parasite densities, the simultaneous amplification of *Poc* and *Pow* DNA in a single reaction tube allows our assay to have shorter turnaround time and require less materials compared to published singleplex assays [17, 18, 44]. The duplex assay’s high specificity, short turnaround time and capacity for high-throughput use make it an especially useful tool for field studies of symptomatic *P. ovale* infection.

The prevalence of *P. ovale* infections in symptomatic individuals from the published study was 6.4% [25], while lower prevalence of *P. ovale* infections were found in nationally representative, asymptomatic adults (0.5%) and children (4.7%) in DRC [28]. *P. ovale* spp. infections in symptomatic individuals had significant association with rural residence and co-infection with *P. falciparum*, which is consistent with previous studies [28, 37, 47]. Also similar to the results of previous studies [48, 49], bed net use was found to be protective against *P. ovale* malaria in our study, indicating that it is an effective strategy to reduce *P. ovale*-related malaria burden. People who reported malaria infection within six months before enrollment showed higher odds of *P. ovale*-related malaria infection; this finding could be attributed to re-infection, inadequate treatment of previous malaria infections, or *P. ovale* relapse [50]. Existing evidence indicates that the most prevalent *P. ovale* spp. vary across different countries [51]. Using the present duplex qPCR assay, we detected both *Poc* and *Pow* across three provinces in the DRC, with *Poc* more prevalent in symptomatic individuals. A previous, nationally representative study in asymptomatic and symptomatic school-age children in DRC also showed high prevalence of *Poc* [47].

Several limitations of our approach should be highlighted. First, the assay’s relatively low sensitivity at lower parasite densities, particularly for *Pow* detection, limits its utility in low-density or asymptomatic infections. Thus, the prevalence of *Poc* and particularly *Pow* in our study are likely underestimated. This limitation could be overcome in the future by combining a pan-*P. ovale* spp. 18S rRNA assay (e.g. such as that used by Mitchell et al. [2]) with our *Poc lsa3* assay, allowing definitive identification of *Poc* (pan-*P. ovale* spp.-positive, *Poc lsa3-*positive) and deductive identification of *Pow* mono-infection (pan-*P. ovale* spp.-positive, *Poc lsa3-*negative).

Second, our *Poc* and *Pow* prevalence estimates are derived from people presenting with malaria symptoms to health facilities in three DRC provinces. The DRC is a large and diverse country. Our results provide further insight into the epidemiology of *Poc* and *Pow* in three large regions but do not provide precise, nationally representative estimates, nor information about asymptomatic infections. Third, crude associations were analyzed in this study to explore possible factors related to *P. ovale*; no causal relationships were assessed. Fourth, the assay targets two non-essential genomic regions at risk of deletion or disruption if future treatment choices are tied to diagnosis, as has been proposed for *P. falciparum* and observed for *Chlamydia trachomatis* non-essential diagnostic targets [52, 53]. However, this hypothetical threat is unlikely to be realized any time soon. Malaria programs in Africa focus largely on *P. falciparum* and do not routinely offer radical cure to clear *P. ovale* spp. hypnozoites.

In conclusion, we developed and validated a novel, highly specific duplex real-time PCR assay capable of detection and differentiation of *Poc* and *Pow*. Though our assay’s sensitivity at lower parasite densities could be further improved, its streamlined work-flow reduces complexity and is well suited for high-throughput use in field studies. As some countries progress toward malaria elimination, improved assays for *Poc* and *Pow* like that presented here will become more important and open the way to improved understanding of *P. ovale* spp. epidemiology, clinical impact, to inform eradication strategies.

## Data Availability

All data and related metadata underlying reported findings have been provided as part of the submitted article.

## Acknowledgements

We thank the study teams and participants in the DRC and Tanzania research studies from which samples were derived. The following reagents were obtained through BEI Resources, NIAID, NIH: diagnostic plasmid containing the small subunit ribosomal RNA gene (18S) from *Plasmodium ovale*, MRA-180, contributed by Peter A. Zimmerman.

## Author contributions

J.B.P. and W.H. conceptualized the study. J.B.P., J.J.J., J.T.L., and W.H. designed experiments. M.K., A.T., F.P., A.K., and B.N. collected field samples. W.H., V.R.P., K.L.T., and R.S. performed laboratory and epidemiological analyses. W.H. prepared the figures and wrote the first draft. All authors reviewed and approved the final draft.

## Supporting information

**S1 Table. Candidate primer and probe sets evaluated for the detection of *P. ovale curtisi* and *P. ovale wallikeri*.**

**S2 Table. Limit of detection of the optimized, duplex *P. ovale curtisi* and *P. ovale wallikeri* assay versus serially diluted plasmid DNA.**

**S1 Fig. 95% lower limits of detection determined using probit analysis.** A) *P. ovale curtisi* 95% lower limits of detection (4.2 parasites/µl [95% CI 3.1-9.5]). B) *P. ovale wallikeri* 95% lower limits of detection (41.2 parasites/µl [95% CI 33.3-58.3]). Confidence intervals are shown in lighter shade.

